# Exploring proteins as intermediates between physical activity and cancer in UK biobank

**DOI:** 10.64898/2026.09.10.26362738

**Authors:** Lucy J. Goudswaard, Matthew A. Lee, Lisa M. Hobson, Rebecca C. Richmond, Emma E. Vincent, Richard M. Martin, Marc Gunter, Brigid M. Lynch, Kostas K. Tsilidis, Sarah J. Lewis

## Abstract

**Introduction:** Higher physical activity (PA) is associated with lower risk of several cancers, but the biological mechanisms underlying these associations remain unclear. We integrated observational and Mendelian randomization (MR) approaches to investigate circulating proteins as potential intermediates linking PA to cancer risk.

**Methods:** We analysed accelerometer-derived PA measures (∼100,000 participants), circulating protein levels (∼53,000 participants) derived using the Olink Explore 3072 panel and cancer incidence (breast, colorectal, endometrial, and prostate cancers) via linked registry data among UK Biobank (UKB) participants. Observational associations were assessed using regression models and Cox proportional hazard models adjusting for demographic, lifestyle, and clinical covariates, including body mass index (BMI). Proteins associated with both overall acceleration average and cancer incidence were taken forward in a two-sample and two-step MR analyses using genome-wide association data.

**Results:** Higher overall acceleration average was observationally associated with lower risk of breast, colorectal, and endometrial cancer, with attenuation after BMI adjustment for endometrial and colorectal cancer, and a positive association with prostate cancer. MR analyses supported a protective causal effect of higher overall PA on breast, colorectal, and prostate cancer risk, and suggested a much larger effect than observational analyses. Overall, PA was observationally associated with 436 circulating proteins of which 44 were also associated with incident cancer risk. MR analyses provided some support that TNFSRF13B was influenced by physical activity in the same direction as the observational analyses but found conflicting evidence for an effect of TNFSRF13B on breast cancer, although the MR analysis is consistent with a protective effect of physical activity on breast cancer risk.

**Conclusions:** While higher overall physical activity appears to causally reduce risk of several cancers, circulating proteins associated with PA largely do not show strong evidence of being intermediates of these effects, suggesting they may act primarily as biomarkers of physical activity rather than causal intermediates.

## Introduction

Higher levels of physical activity (PA) have consistently been shown to be associated with a reduced risk of several cancers, including endometrial, colorectal and breast cancer (1, 2). However, evidence for prostate cancer has been more complex: while some studies have demonstrated a positive association between PA and prostate cancer, this finding is reported to be partially attributed to detection bias where men who are more physically active may be more health-conscious and therefore more likely to undergo prostate-specific antigen (PSA) testing leading to greater detection rates (2). Additionally, evidence also suggests that increased physical activity following a prostate cancer diagnosis is associated with improved survival (3); however, this finding may be at least partly explained by reverse causation and confounding, whereby diagnosis encourages behavioural change among those who are still physically fit and who have a better prognosis.

Mendelian randomization (MR) studies, which use genetic variants as proxies for PA have provided evidence for a potentially protective causal effect of higher levels of accelerometery-derived PA on the risk of colorectal, breast and endometrial cancers and, in contrast to observational studies, a protective effect on overall prostate cancer risk (4–7).

These findings provide complementary evidence of the importance of PA as a modifiable risk factor for cancer development. Despite this evidence, the molecular intermediates which link PA to cancer risk are not fully understood. Characterising these biological pathways could inform strategies to optimise PA for cancer prevention and provide protein targets for potential modulation. Advances in high throughput proteomic profiling from small volumes of blood now offer opportunities to identify biological pathways influenced by PA that may underlie its protective effects on cancer. In particular, the recent availability of plasma proteins in over 50,000 individuals in UK Biobank (UKB) has enabled well-powered studies into the role of circulating proteins in health and disease (8).

In this study, we combined observational and MR approaches to investigate associations between accelerometer-measured PA, circulating proteins levels and cancer risk, aiming to investigate potential molecular intermediates that may help explain the link between PA and cancer development.

## Methods

### Study overview

This study used a combination of observational and MR approaches to further understand the associations between physical activity, circulating protein levels and cancer risk (**Figure 1**).

The main objectives were:

1. A) Assess the associations between accelerometer-measured physical activity and circulating protein levels in the UK Biobank. B) Examine the relationships between physical activity and incident breast, prostate, endometrial, and colorectal cancers. C) Evaluate the associations between physical activity–related proteins and the risks of breast, prostate, endometrial, and colorectal cancer.
2. For proteins associated with overall acceleration average and cancer outcomes in a directionally concordant manner, apply two-step MR to investigate potential causal relationships.

### UK Biobank (UKB)

The UKB is a prospective study which recruited 500,000 participants across 22 assessment centres across the United Kingdom. At initial recruitment in 2006-2010, participants were aged 40-69 years (9). This study has collected information about participants using a combination of questionnaires, physical and clinical measures (9). UK Biobank also performs centralised linkage of participants to national hospital, cancer, and death registry records.

### Physical activity variables

Physical activity measurements were derived using an Axivity AX3 wrist-worn triaxial accelerometer. Around 100,000 UKB participants wore the accelerometer on their dominant wrist for seven days between 2013 and 2015 (10). Three derived physical activity variables were used for this study: overall acceleration average (measured in milli-gravities), which gives the average acceleration across the seven days (and has been validated with using metabolic equivalent tasks - METs); moderate-vigorous PA, which indicates the proportion of time spent doing moderate-vigorous activities; and a sedentary variable, which indicates the proportion of time spent sedentary (10, 11). Participants whose data could not be calibrated or who did not wear the device for the recommended time were excluded.

Extreme outliers (± 5 SDs from the mean) were excluded. Overall acceleration average and sedentary behaviour variables were inverse rank normal transformed using the rntransform() function from the “moosefun” R package. Moderate to vigorous activity was transformed into quartiles due to the large number of zero values.

### Proteomics

Proteins were measured in UKB, using the Olink Explore 3072 panel, from 53,016 participant blood samples collected at baseline (2006–2010). Olink uses the proximity extension assay (PEA) using a dual-antibody approach to detect and quantify proteins in normalised protein expression units (NPX) on a log2 scale (12). A total of 2923 unique proteins were detected across individuals (8). Protein levels were inverse rank normal transformed due to there being distributions that were not normal based on a Shapiro-Wilk W statistic of < 0.95. This transformation also matches pre-processing of the proteomic data during the GWAS used in MR (described later in this manuscript).

### Cancer registry data

Cancer cases were defined by ICD-10 or ICD-9 codes and were acquired from a variety of sources such as hospitals, cancer centres and general practices, with further information provided by UKB (13) (14). Instances are defined as a different timepoint where the cancer registry information was integrated into the dataset to capture newly diagnosed cancer cases, typically occurring every 6-12 months (14). There were 22 ICD-10 instances, and15 ICD-9 instances that were used in the current analysis, with further details of dates of diagnoses provided elsewhere (15). The cancers of interest were breast cancer (female only; ICD-10 C50, C50.0, C50.1, C50.2, C50.3, C50.4, C50.5, C50.6, C50.7, C50.8, C50.9, ICD-9 174, 174.0, 174.1, 174.2, 174.3, 174.4, 174.5, 174.6, 174.7, 174.8, 174.9), endometrial cancer (ICD-10 C54, C54.1, C54.2, C54.3, C54.9, C55, ICD-9 179, 1799, 180, 182, 1820, 1821, 1828), prostate cancer (ICD-10 C61, ICD-9 185) and colorectal cancer (ICD-10 codes C18, C18.0, C18.1, C18.2, C18.3, C18.4, C18.5, C18.6, C18.7, C18.8, C18.9, C19, C20, ICD-9 1530, 1531, 1532, 1533, 1534, 1535, 1536, 1537, 1538, 1539, 1540, 1541, 1542, 1543, 1548).

### Covariates

We adjusted for variables plausibly having an effect on physical activity measures, circulating proteins or cancer risk (16). Covariates included; age at enrolment, sex, study centre, fasting status at blood collection and educational level. Smoking was defined as ever/never, alcohol status as current/previous/never. Educational level was grouped into an ordered variable: (no qualifications (0); CSEs/O levels/GSCEs (1); A levels/AS levels (2); NVQ/HND/HNC (3); other profession qualifications e.g. nursing, teaching (4); college or university degree (5). In separate models, we additionally adjusted for body mass index (BMI) to explore possible biological pathways that occur independent of BMI.

### Statistical analyses

Individuals who elected to withdraw from UKB were excluded from analyses based on the withdrawals list on 17^th^ December 2024. Participants were included in analyses if they either had (1) both PA and proteomics data or (2) PA and cancer outcome data or (3) proteomics and cancer data (with information on covariates). There were 127,474 participants included in at least one analysis, but for each specific analysis we present details of the number of participants included.

### Associations between physical activity and circulating proteins

As accelerometery was measured after blood sample collection (which was used to determine circulating protein levels), our analyses were performed under the assumption that physical activity is generally stable within individuals and a person’s activity in 2013-2015 generally reflects an individual’s activity levels between 2006-2010. However, we were unable to remove the effect of reverse causation in this analysis. Measurement of physical activity using accelerometery was only performed once. To test our assumption that physical activity is likely to be consistent across this timeframe, we explored the stability of self-reported physical activity across instances, using questionnaire data. Specifically, we used MET minutes per week for all activity; this was measured in participants at baseline (instance 0) and then again from 2014 onwards (instance 2). There was a moderate positive correlation across instance 0 (2006–2010) and instance 2 (2014+) (N=14,710, r = 0.54). We explored the association between physical activity measures (overall acceleration average, sedentary behaviour, moderate to vigorous activity) and circulating proteins using linear regression “lm()”, with physical activity as the exposure and circulating protein levels as continuous outcomes. Linear regression was performed as three models. The models were: (1) adjustment for age and sex only, (2) adjustment for age, sex, smoking, alcohol, study centre, fasting status, education, (3) adjustment for age, sex, smoking, alcohol, study centre, fasting status, education, body mass index.

We performed a Bonferroni correction, taking into account the estimated number of independent proteins using a correlation cut-off of Spearman’s rho of r=0.5 (17), therefore the p-value for guiding associations was 0.05/1885=2.6×10^−5^.

### Associations between physical activity and cancer risk

Associations between physical activity exposures and incident cancer risk were examined using both logistic regression and Cox proportional hazards models, each fitted with two levels of covariate adjustment. Logistic regression models were used to estimate odds ratios (ORs) for incident cancer occurring after the 31st December 2015 cut-off. Cox proportional hazards models were used to estimate hazard ratios (HRs), with a 2-year delayed entry from baseline to reduce potential reverse causation. In both modelling frameworks, level 1 adjustment included age at recruitment, sex, smoking status, alcohol intake, and education, and level 2 additionally included body mass index (BMI). Cox models were stratified by study centre. Follow-up time in Cox models was censored at the earliest of cancer diagnosis, death, or administrative censoring (9th May 2022). Sex-specific analyses were applied where appropriate (prostate cancer in males; breast and endometrial cancer in females).

### Associations between circulating proteins and cancer risk

For proteins associated with overall physical activity, we further investigated their associations with incident cancer risk using Cox proportional hazards models. Time-at-risk was defined using age as the underlying time scale, with left truncation implemented via delayed entry 2 years after blood draw. Models were stratified by study centre and adjusted for age at recruitment, sex, smoking status, alcohol intake status, educational attainment, and body mass index. Participants were censored at the earliest of cancer diagnosis, death, or administrative censoring (9th May 2022). Individuals with missing exposure or covariate data were excluded from the analyses, and cancer-specific sex restrictions were applied where appropriate. Here, we adjusted for the number of proteins tested, using a multiple testing threshold of 0.05/436 = 1.1×10^−4^ to infer strong evidence for an effect. However, as this is as discovery analysis, proteins which were associated with cancer at p<0.001 were taken forward for MR analysis.

### Functional annotations

We performed separate lookups of the proteins we identified as being upregulated and downregulated in response to overall acceleration average to determine which biological pathways were represented using STRING (https://string-db.org/).

### Mendelian randomization analyses Overview

We conducted a series of bidirectional two-sample Mendelian randomization (MR) analyses to explore potential causal relationships between overall acceleration average (PA), circulating proteins, and cancer risk. MR analyses are based on three key instrumental variable assumptions. (i) Relevance: the selected genetic variants are strongly associated with the exposure. (ii) Independence: the genetic variants are not associated with confounding factors that influence both the exposure and the outcome. (iii) Exclusion restriction: the genetic variants affect the outcome exclusively through the exposure, with no alternative pathways (i.e., no horizontal pleiotropy).

Proteins included in these analyses were brought forward from observational analyses, such that proteins which had evidence for an association with overall physical activity and with any of the cancer outcomes in fully adjusted models were included; therefore for these analyses we were looking for consistency with the observational analyses rather than applying strict Bonferroni corrections. First, we estimated the causal effect of overall PA on circulating protein levels. Second, we estimated the causal effect of PA on cancer outcomes. Third, we estimated the effect of the circulating protein on cancer risk. Finally, we explored these relationships in the reverse direction to evaluate the possibility of reverse causality.

### Genetic instruments for physical activity

In our Mendelian randomization analyses we used accelerometer-derived overall physical activity as our exposure variable due to this being the variable which had the strongest relationship with cancer incidence. To select our instruments for overall activity we used two previously published GWAS, both performed on UKB data.

The first GWAS was performed by Klimentidis et al. in 91,084 participants in UKB (18). An adjusted overall acceleration average phenotype was derived after accounting for age, sex, genotyping chip, first ten genomic principal components, centre, season (month) of centre visit, with the GWAS performed with BOLT-LMM software (19).

The second was performed by Doherty et al. (20) on the overall acceleration average derived from accelerometery, which included 91,105 individuals of European descent. They also used BOLT-LMM software (19), accounting for related individuals and those of varying genetic ancestry. The model adjusted for assessment centre, genotyping array, age, age squared and season of accelerometer wear.

The two overall physical activity GWAS were measured in different units: in the GWAS by Klimentidis et al. (main analysis), the GWAS betas were measured in milligravities, and for the GWAS by Doherty et al. (sensitivity analysis), the GWAS betas were converted to standard deviation (SD) units (18, 20). For comparison with the observational analyses and with the analyses using the genetic instruments from the GWAS by Doherty et al., we converted the results from the MR analyses using the Klimentidis et al. GWAS to per SD units of physical activity, by multiplying the betas by the SD.

Single nucleotide polymorphisms (SNPs) strongly (p<5×10^−8^) and independently (R^2^<0.001) associated with overall acceleration were used as genetic instruments (18, 20).

### Protein genetic instruments

Circulating protein GWAS were performed by Sun et al. (8). Here, proteins were measured in EDTA plasma in 34,557 participants of European ancestry from UKB using the Olink Explore 3072 panel. Prior to genome-wide analysis, protein values measured in NPX units on the log2 scale (21) were inverse rank normal transformed. Following this, a whole-genome regression model using a leave one chromosome out scheme was performed with REGENIE (version 2.2.1). This model was adjusted for age, age^2^, sex, age x sex, age^2^ x sex, batch, centre, genetic array, time between blood sampling measurements and the first 20 principal components. For each protein, SNPs were identified from the protein-coding gene region (including a 1 Mb window) using a p-value threshold of 5×10^−8^ and an LD independence of R^2^ < 0.001. Where no SNP met this criteria, a single SNP was identified using approximate Bayes factors (22) (23). Several proteins had multiple measurements in UK biobank as they were measured on more than one of Olink’s panels, for example Cardiometabolic I and Inflammation I. For these proteins the following Olink IDs were used: tumour necrosis factor – OID20074, interleukin-6 - OID20101, leiomodin-1 - OID30212.

### Cancer genetic instruments

Genome-wide association study data from cancer consortia were used for the MR analyses of colorectal, breast, prostate, endometrial cancers (24–27). Studies were restricted to those that included individuals of European ancestry. A brief summary of these GWAS is provided below, with more information in the **Supplementary materials** and full methodological details are available in the original publications.

For the prostate cancer GWAS, there were 79148 cases and 61106 controls (OpenGWAS ID: ieu-b-85) from the PRACTICAL consortium (24).

Breast cancer summary statistics were obtained from the Breast Cancer Association Consortium (BCAC) OncoArray meta-analysis (25) (Open GWAS ID: ieu-a-1126). This meta-analysis combined data from the OncoArray, iCOGS and 11 previous GWAS, totalling 122,977 cases and 105,974 controls.

GWAS summary statistics for endometrial cancer were generated with 12,906 cases and 108,979 controls 13 studies of endometrial cancer (27).

The colorectal cancer GWAS was performed using studies from the Genetics and Epidemiology of Colorectal Cancer Consortium (GECCO), the Colorectal Cancer Transdisciplinary Study (CORECT) and the Colon Cancer Family Registry (CCFR). The GWAS included 52,775 colorectal cancer cases and 45,940 controls (26).

Germline genetic variants that were strongly (p<5×10^−8^) and independently (R^2^ < 0.001) associated with cancer risk were used to instrument cancer risk (24–27).

### Mendelian randomization methods

Two-sample Mendelian randomization was performed using the “TwoSampleMR” R package –(28). Where ≥3 SNPs were available, an inverse variance weighted (IVW) method with random effects was used; where 2 or more SNPs were available, an IVW method with fixed effects was used; where 1 SNP was available, the Wald ratio was used (29–31). Sensitivity analyses (where ≥3 SNPs were available) included weighted median, weighted mode and MR Egger, which were applied to assess the robustness of the causal estimates and to detect potential violations of MR exclusion assumption, which can be caused by horizontal pleiotropy (32–34). Heterogeneity was explored through use of Cochran’s Q statistic (35). To assess the relevance assumption, we calculated instrument F-statistics, to assess independence we reported other phenotypes associated with our instruments based on data from GWAS catalog https://www.ebi.ac.uk/gwas/. Results are presented as an SD change (continuous) or odds ratio (dichotomous) per SD change in the exposure. Strong evidence for effect of PA on circulating protein and circulating protein on cancer outcome analyses was considered using a Bonferroni-adjusted p-value of 0.05/44 = 0.001.

### Sample overlap

Sample overlap can result in weak instrument bias within MR studies, with the degree of weak instrument bias induced by sample overlap being proportional to the degree of overlap (36). Some overlap existed between the GWAS used for the exposures and the outcome, for physical activity and circulating proteins, both of which included UK Biobank participants, the degree of sample overlap was limited; For example, based on exploring overlap of the individual level data in UKB, approximately 20% of participants in the protein GWAS were also included in the physical activity GWAS, and about 12% of the physical activity GWAS sample overlapped with the protein GWAS sample (36).

## Results

### Observational analyses in UKB

**Table 1** displays the summary of participants included in the study. Study individuals were 55% female, the mean BMI was 26.9 kg/m^2^ (SD 4.6 kg/m^2^), and the majority of participants were ever smokers and at the time of study most (94%) consumed alcohol.

**Table 1.** Participant characteristics for those included in the observational main analysis (N up to127,474)

| Characteristic | N | Mean (SD) |
| --- | --- | --- |
|  |  | or<br>n / N % |
| Age at baseline (years) | 127,474 | 56 (8) |
| Sex | 127,474 |  |
| Female |  | 70,076 / 127474 (55%) |
| Male |  | 57,398 / 127474 (45%) |
| Education | 127,160 |  |
| 0 |  | 13,170 / 127160 (10%) |
| 1 |  | 19,507 / 127160 (15%) |
| 2 |  | 7,721 / 127160 (6.1%) |
| 3 |  | 15,139 / 127160 (12%) |
| 4 |  | 19,708 / 127160 (15%) |
| 5 |  | 51,915 / 127160 (41%) |
| Body mass index (kg/m<sup>2</sup>) | 127,297 | 26.9 (4.6) |
| Ever smoker | 127,369 | 76,661 / 127369 (60%) |
| Alcohol intake | 127,443 |  |
| Never |  | 4,182 / 127443 (3.3%) |
| Previous |  | 3,851 / 127443 (3.0%) |
| Current |  | 119,410 / 127443 (94%) |
| Overall acceleration average (milligravities) | 95,183 | 28.0 (8.0) |
| Moderate to vigorous activity (proportion) | 95,136 | 0.028 (0.023) |
| Sedentary behaviour (proportion) | 95,305 | 0.39 (0.08) |

### Association between physical activity and circulating proteins

In our age and sex adjusted analysis, 922 (31.5%) out of the 2923 proteins passed our Bonferroni test for association with overall physical activity **(Supplementary Table 1)**. There were 436 (14.8%) proteins associated with overall physical activity when adjusting for confounders including body mass index **(Supplementary Table 3)**. Of these, 114 proteins were also associated with both moderate to vigorous activity and sedentary behaviour; 120 were also associated with moderate to vigorous physical activity only; 43 were also associated with sedentary behaviour; and 159 were not associated with either moderate to vigorous physical activity or sedentary behaviour (**Figure 2** and **Supplementary Tables 1-9**). The proteins most strongly positively associated with overall physical activity in our fully adjusted model were three integrins: integrin subunit alpha v (ITGAV, beta = 0.33 SD, 95% CI 0.31 to 0.35, p=8.87 ×10^−245^), integrin alpha M chain (ITGAM, beta = 0.26, 95% CI 0.24 to 0.28, p=6.40 ×10^−139^), and integrin subunit alpha 11 (ITGA11, beta = 0.24 SD, 95% CI 0.22 to 0.26, p=1.82 ×10^−120^). The protein which had the strongest inverse association with overall physical activity was leptin (LEP, beta = -0.12 SD, 95% CI -0.13 to -0.11, p=2.98×10^−89^). Of the 436 proteins associated with overall physical activity in our adjusted analysis, 310 were found at lower levels with increasing activity. Biological processes represented within the downregulated proteins were: response to stress (114 proteins), response to external stimuli (110 proteins), immune system processes (105 proteins), cell surface signalling (102 proteins), and cell adhesion (49 proteins) (https://string-db.org/). Among the 126 proteins which were found at higher levels with increasing activity, the following biological processes were represented: response to stimuli (85 proteins), multicellular organismal processes (84 proteins), anatomical structure development (70 proteins), cell surface receptor signalling (43 proteins), cell adhesion (40 proteins), cell motility (29 proteins) (https://string-db.org/).

### Association between physical activity and cancer risk

The association between physical activity and cancer risk was explored using logistic regression and Cox PH models (**Figure 3**). For incident cancer in the primary logistic regression model there were 604 colorectal, 918 breast, 1283 prostate and 160 endometrial cancer cases, while the Cox PH model included fewer incident cases due to the two-year time lag. Further details about the number of cases and controls are in **Supplementary Table 10** and **Supplementary Table 11.** In general, there was good agreement between the two models. After adjusting for covariates (but before adjustment for BMI), one normalised SD increase in overall acceleration average was associated with lower risk of colorectal cancer (logistic regression OR 0.92, 95% CI 0.85-1.01, p=0.065; Cox PH HR 0.93, 95% CI, 0.85-1.02, p=0.11), a protective effect on breast cancer (logistic regression OR 0.92, 95% CI 0.86-0.99, p=0.02; Cox PH HR 0.93, 95% CI 0.86-1.00, p=0.07) and similarly a protective effect on endometrial cancer risk (logistic regression OR 0.80, 95% CI 0.68-0.94, p=7.8×10^−3^; Cox PH HR 0.84,95% CI 0.70-0.99, p=0.047). The estimates for the association between overall physical activity and risk of prostate cancer were both positive (logistic regression OR 1.07, 95% CI 1.01-1.14, p=0.01; Cox PH HR 1.05, 95% CI 0.99-1.12, p=0.01).

After adjustment for BMI, the effects of overall physical activity on breast cancer (logistic regression OR 0.93, 95% CI 0.86-1.00, p=0.05; Cox PH HR 0.94, 95% CI 0.87-1.02, p=0.0.13) and prostate cancer (logistic regression OR 1.07, 95% CI 1.01-1.13, p=0.03; Cox PH HR 1.04, 95% CI 0.98-1.11, p=0.18) were largely unchanged. Whereas for colorectal cancer, the BMI-adjusted estimates were attenuated slightly in both models, although the Cox model still showed some weak evidence of an effect (logistic regression OR 0.96, 95% CI 0.88-1.05, p=0.39; Cox PH HR 0.96, 95% CI 0.88-01.06, p=0.45). For endometrial cancer, the protective association of overall physical activity attenuated after BMI adjustment in both models (logistic regression OR 0.97, 95% CI 0.81-1.16, p=0.75; Cox PH HR 1.00, 95% CI 0.83-1.21, p=0.98).

Moderate to vigorous activity and sedentary behaviour displayed broadly similar associations with cancer risk compared to overall acceleration average in terms of directions of effect, but effects were generally weaker. Although there was no evidence of an effect of moderate to vigorous activity on colorectal cancer risk and even a suggestion of an effect in a positive direction.

### Association between circulating proteins and cancer risk

Among the 436 proteins observationally associated with overall physical activity, 28 protein-incident cancer associations, representing 25 unique proteins, met the Bonferroni-adjusted threshold after adjustment for covariates including BMI (**Supplementary Table 12**). Most of these associations were with breast cancer. The strongest were observed for lysosome-like protein 2 (LYZL2) and cadherin EGF LAG seven-pass G-type receptor 2 (CELSR2), with higher circulating concentrations of both proteins associated with a higher risk of breast cancer.

Some proteins were associated with more than one cancer type. For example, higher amphiregulin (AREG) concentrations were associated with a higher risk of both breast cancer (HR 1.16, 95% CI 1.11-1.22; p = 3.8 × 10⁻□) and colorectal cancer (HR 1.24, 95% CI 1.14-1.34; p = 1.8 × 10⁻□). A further 21 associations, representing 21 unique proteins, showed suggestive evidence of association, defined as 1.1 × 10⁻□ ≤ p < 0.001. Because two proteins had associations falling in both the Bonferroni-adjusted and suggestive categories for different cancer types, the 49 associations involved 44 unique proteins overall. All 44 proteins showing either Bonferroni-significant or suggestive evidence of association with at least one cancer are presented in **Figure 3**.

### Mendelian randomization analyses Physical activity and cancer risk

The instruments used for our primary overall physical activity analysis were rs55657917 and rs59499656 (18); these had F statistics of 56 and 36, respectively. The former SNP has been associated with mood instability (37) and the latter with high density lipoprotein cholesterol levels and body fat percentage (38). In the secondary analysis five instruments derived by Doherty et al (20) with instrument F statistics ranging from 27-44, were used. One SNP rs59499656 was represented in both instruments, the other SNP rs55657917 identified in the GWAS by Klimentidis was within 2.6 megabases of a SNP identified in the Doherty et al. GWAS (20). Within the Doherty et al. instrument rs6775319 has also been associated with C-reactive protein levels, fruit-taste liking and whole body fat mass and rs564819152 has been associated with various brain measurements, body fat, BMI and red blood cell diameter, whereas, rs6895232 and rs2696625 have only been associated with physical activity (GWAS catalog - https://www.ebi.ac.uk/gwas/). These instruments and their association with overall acceleration average are presented in **Supplementary Table 13.**

The MR estimates for the effect of overall physical activity on cancer risk, using the Klimentidis instrument [18], indicated that higher levels of overall physical activity lowered the risk of colorectal cancer (OR 0.59 per SD higher physical activity, 95% 0.37 to 0.94, p=0.03), breast cancer (OR 0.42, 95% CI 0.31 to 0.58, p=3.1×10^−8^) and prostate cancer (OR 0.48, 95% CI 0.33 to 0.72, p=2.9×10^−4^), but there was weaker evidence of an effect on endometrial cancer risk (OR 0.75, 95% CI 0.36 to 1.56, p=0.44). These estimates are displayed in **Figure 5** and in **Supplementary Table 14**). Effects were consistent in direction using alternative instruments by Doherty et al. for overall physical activity (20), and across other MR methods (**Supplementary Table 14**). In the reverse direction, there was no evidence for an effect of genetic liability to cancer risk having an effect on physical activity (**Supplementary Table 15)**.

### Physical activity and circulating proteins

The 44 unique proteins associated with both PA and at least one cancer outcome in observational analyses were taken forward into Mendelian randomization analyses. We first explored the effect of overall physical activity on this set of circulating proteins, using the 2 SNP instrument derived from the Klimentidis et al. GWAS as the primary analysis. The MR estimates using our primary instrument are presented in **Figure 6A** and estimates using both our primary and alternative instruments are provided in **Supplementary Table 16.** There was evidence (p<0.001) for an effect of physical activity on 2 proteins. There was strong evidence from our MR analyses that physical activity reduced levels of tumour necrosis factor receptor superfamily member 13B (TNFRSF13B), while raising levels of cathepsin O (CTSO). In the reverse MR (effect of protein on physical activity), there was some evidence that higher levels of CTSO may influence physical activity levels, however this estimate did not pass multiple testing (**Supplementary Table 17**). Of these two proteins, the MR estimate was only in a consistent direction with the observational analyses for TNFRSF13B. In addition, trefoil factor 3 (TTF3) levels were reduced with increasing physical activity in both observational and MR analyses using the Doherty et al. instrument.

### Circulating proteins and cancer risk

There were no proteins where MR evidence showed a causal effect on any of the 4 cancer outcomes at a Bonferroni-adjusted p-value. In addition, there was little consistency between the MR and observational analyses for the effect of proteins on cancer risk (**Supplementary Table 18**, **Figure 6B**). For example, the association between TNFRSF13B and breast cancer observationally per SD higher protein was HR 0.86, 95% CI 0.82-0.91, p=4.7×10^−8^, whereas in the MR it was OR 1.17, 95% CI 1.05-1.32, p=6.0×10^−3^. Reverse MR did not suggest that breast cancer risk influences TNFRSF13B (**Supplementary Table 19**). Results of heterogeneity tests for all MR analyses are presented in **Supplementary Table 20**.

## Discussion

In this study we explored the relationship between physical activity, circulating proteins and four common cancers using observational and Mendelian randomization approaches.

Consistent with the literature, we observed inverse observational associations between overall physical activity and risk of breast, colorectal and endometrial cancer, with attenuation for endometrial cancer after adjusting for BMI, likely reflecting mediation by adiposity (39). The positive estimate for the observational association between physical activity and prostate cancer risk has also been observed in other studies (39), and is thought to, in-part, reflect greater screening uptake among more health-conscious and physically active men (2). We found MR evidence that higher overall physical activity reduced risk of breast, colorectal, and prostate cancer, supporting potential causal effects that have previously been reported (4–6), The estimates from our MR analysis suggested much greater protective effects of overall physical activity than was seen in our observational analyses and effects were consistent across the two different instruments we used in our MR analysis. The difference in magnitude of effect between our observation and MR analyses may have arisen because we are not capturing lifetime exposure or exposure during a critical period in our observational analyses, or because our MR estimates are biased. It is possible that confounding is present in our MR analyses. However, if this is the case, it is unclear what confounding factor could exert such strong effects across breast, colorectal, and prostate cancer. Adiposity is unlikely to explain this pattern, as higher BMI has consistently been shown to decrease overall breast and prostate cancer risk and increase risk of endometrial and colorectal cancer in MR studies (40, 41). Unlike previous studies, we did not find strong evidence for a causal effect of overall physical activity on endometrial cancer, however this could be due to a different SNP selection (7).

In our fully-adjusted analyses, we identified 436 observational associations between accelerometer-derived physical activity representing 15% of circulating proteins tested. We found strong evidence of an inverse association between overall physical activity and leptin, a hormone with a known role in satiety, even after adjustment for BMI. Other effects observed were positive associations between overall physical activity and integrins (ITGAV, ITGAM, ITGA11). These integrin alpha subunits each dimerize with different beta subunits and have a role in cell adhesion. These may raise in circulation following mechanical stimulation (42), or perhaps by an increase in the release of exosomes (43). A previous study exploring protein associations with physical activity also found observational associations between physical activity and integrins, results consistent with the current study

(44). The effects of overall physical activity on these integrin proteins were so strong they could perhaps be used in future as biomarkers of activity. The same integrins were found to be associated with a decreased risk of colorectal and endometrial cancer in our observational analyses, although integrins were not found to be associated with cancer risk in our MR analyses, which means that a causal effect of these proteins on cancer is unlikely.

Forty-four proteins which were associated with physical activity were also associated with cancer risk in our observational analyses. Among these TNFRSF13B and TFF3 showed strong evidence of inverse effects with physical activity in both observational and Mendelian randomization (MR) analyses (with 1 of the 2 instruments), although the effects in our MR analyses were instrument-specific effects which limited robustness. TNFRSF13B encodes TNF receptor superfamily member 13B (TACI), which is a member of the TNF receptor superfamily, involved in NF-kappa-B signalling. We found TNFRSF13B to be associated with a decreased risk of breast cancer in our observational analyses, but an increased risk in our MR analyses. Recent studies have shown that silencing of TNFRSF13B can lead to the death of breast cancer cells and that TNFSRF13B mRNA levels were associated with breast cancer prognosis (45, 46).

TFF3 is a mucosal repair protein that protects and restores epithelial tissues, in particular in the intestine (47). It has been reported that TFF3 expression is elevated in colorectal cancer cells and may promote tumour progression (48). In our observational analyses, we found weak evidence for a positive association between TFF3 and colorectal cancer risk and stronger evidence for a negative association with prostate cancer risk; however, MR analyses did not support the latter association.

Taken together, the above findings do not support a clear two-step causal pathway linking physical activity to cancer risk at the level of the proteome. Although they do highlight some potential pathways worthy of further exploration, in particular pathways involving TNFRSF13B.

The strengths of our study include the use of large datasets, including the incorporation of large cancer consortia GWAS in our MR analyses, and the triangulation of observational and Mendelian randomization (MR) approaches, which allowed us to test the robustness of our findings and explore potential causal relationships. By conducting analyses both adjusted and unadjusted for BMI, we were able to distinguish effects mediated by BMI, such as the relationship between physical activity and endometrial cancer risk, from direct effects of physical activity independent of BMI. We used objective measures of physical activity, capturing total daily activity rather than relying on self-reported activity or specific exercise types. While reverse causation cannot be ruled out in our observational analyses, because physical activity was measured after baseline blood sampling, our MR analyses are inherently less susceptible to reverse causation because germline genetic variants are fixed at conception and are unaffected by disease status.

There are also some limitations to this study. Although different approaches such as observational analyses and MR were used, both approaches used UKB data. In addition, UKB was used both to determine both the effects of physical activity on proteins and the effects of proteins on cancer in the observational analyses, and was both the discovery and test sample for SNPs associated with physical activity and proteins in our MR analysis.

However, to our knowledge UKB represents the only study with GWAS data of physical activity which is accelerometer measured and the largest GWAS study of proteomics data, thus we were limited by the availability of data. This will also affect the generalisability of our findings to other populations which are not primarily white European. Similarly, we assumed linear models and were looking at the effects of exposures within the normal range we were unable to examine the effects of exposures at the extreme end of the spectrum, nor where we able to assess the importance of timing of exposure in this study. We did not include cancer subtypes or localised versus advanced cancers in our analyses as the data were not available in UKB to examine this and sample sizes would have been very small, however, our previous MR analyses have not found strong evidence of differences by cancer subtype (4, 5). Although our protein lookups using STRING helped identify biological pathways shared by the proteins we detected, we were unable to quantify the degree of enrichment or determine whether these pathways were truly enriched. This limitation arises because STRING uses all possible proteins as the reference background, whereas the proteomics panel used to generate our dataset included only a subset of proteins.

Our study shows robust evidence for a protective effect of physical activity on cancer risk and adds to the body of evidence on this. The study highlights the value of integrating molecular and lifestyle data to explore physical activity–cancer relationships. While we identified protein biomarkers associated with physical activity and cancer, robust evidence for causal proteomic intermediates of the relationship between physical activity and cancer was limited. These results provide a framework for future studies to elucidate underlying mechanisms and underscore the importance of replication in larger cohorts.

## Supporting information

Checklist

## Data availability

UK Biobank individual level data was accessed under the application 16391 titled “Investigating the causal effects of accelerometer measures and body composition on chronic disease”, where Dr Rebecca Richmond is the principal applicant. This study has received GECCO approval to use the colorectal cancer GWAS summary statistics file. Scripts used for analyses in this manuscript are located here: https://github.com/lucygoudswaard/PA_protein_cancer_paper. ChatGPT was used to support with analytical code writing.

## Ethics Statement

This research was conducted using the UK Biobank Resource (application number 16391). UK Biobank has ethical approval from the North West Multi-centre Research Ethics Committee (REC reference 16/NW/0274), which was renewed in 2011, 2016 and 2021 and all participants provided informed consent.

## Funding

Funding for this study was provided by a grant (IIG_FULL_2020_019) obtained from Wereld Kanker Onderzoek Fonds (WKOF) as part of the World Cancer Research Fund International grant programme. LJG, SJL, RCR, EEV and RMM are supported by a Cancer Research UK (C18281/A29019) programme grant (the Integrative Cancer Epidemiology Programme). RCR, LJG, SJL, EEV and LMH are part of the Medical Research Council Integrative Epidemiology Unit at the University of Bristol which is supported by the Medical Research Council and the University of Bristol (grant code MC_UU_00011/1). RMM is also supported by the NIHR Bristol Biomedical Research Centre, which is funded by the NIHR and is a partnership between University Hospitals Bristol and Weston NHS Foundation Trust and the University of Bristol. Department of Health and Social Care disclaimer: the views expressed are those of the author(s) and not necessarily those of the NHS, the NIHR, or the Department of Health and Social Care. RMM is a National Institute for Health Research Senior Investigator (NIHR305892). EEV is also supported by the World Cancer Research Fund (funding for IIG_FULL_2024_029 was obtained from World Cancer Research Fund (WCRF), as part of the World Cancer Research Fund International grant programme). LMH is supported in part by grant MR/W006308/1 for the GW4 BIOMED MRC DTP, awarded to the Universities of Bath, Bristol, Cardiff and Exeter from the Medical Research Council (MRC)/UKRI.

## Acknowledgements

We would like to thank Laura Corbin for her discussion and her advice in relation to this manuscript. We thank all participants without whom this research would not have been possible.

## Disclaimer

Where authors are identified as personnel of the International Agency for Research on Cancer/World Health Organization, the authors alone are responsible for the views expressed in this article and they do not necessarily represent the decisions, policy, or views of the International Agency for Research on Cancer/World Health Organization. All authors declare no conflicts of interest.

---

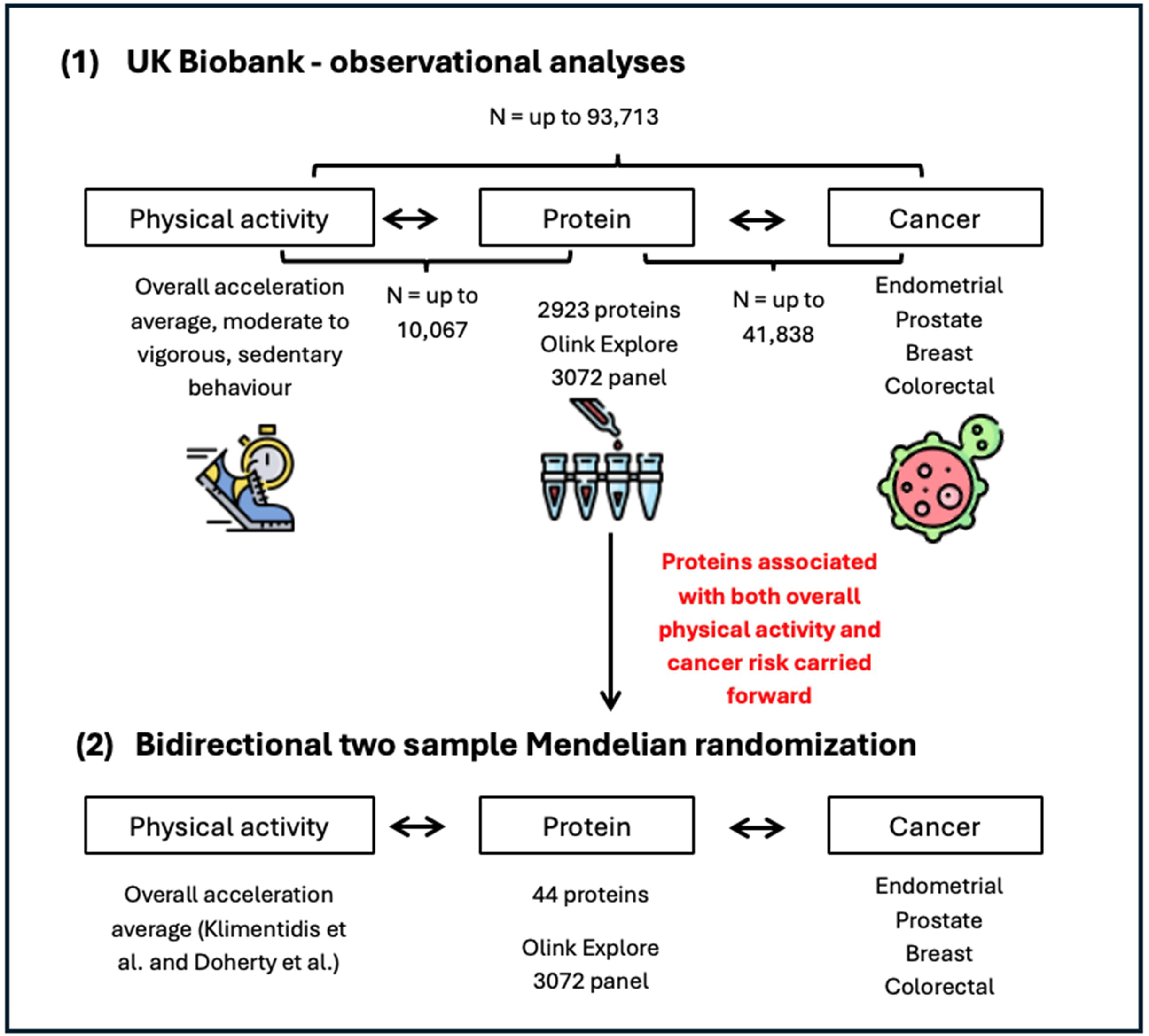

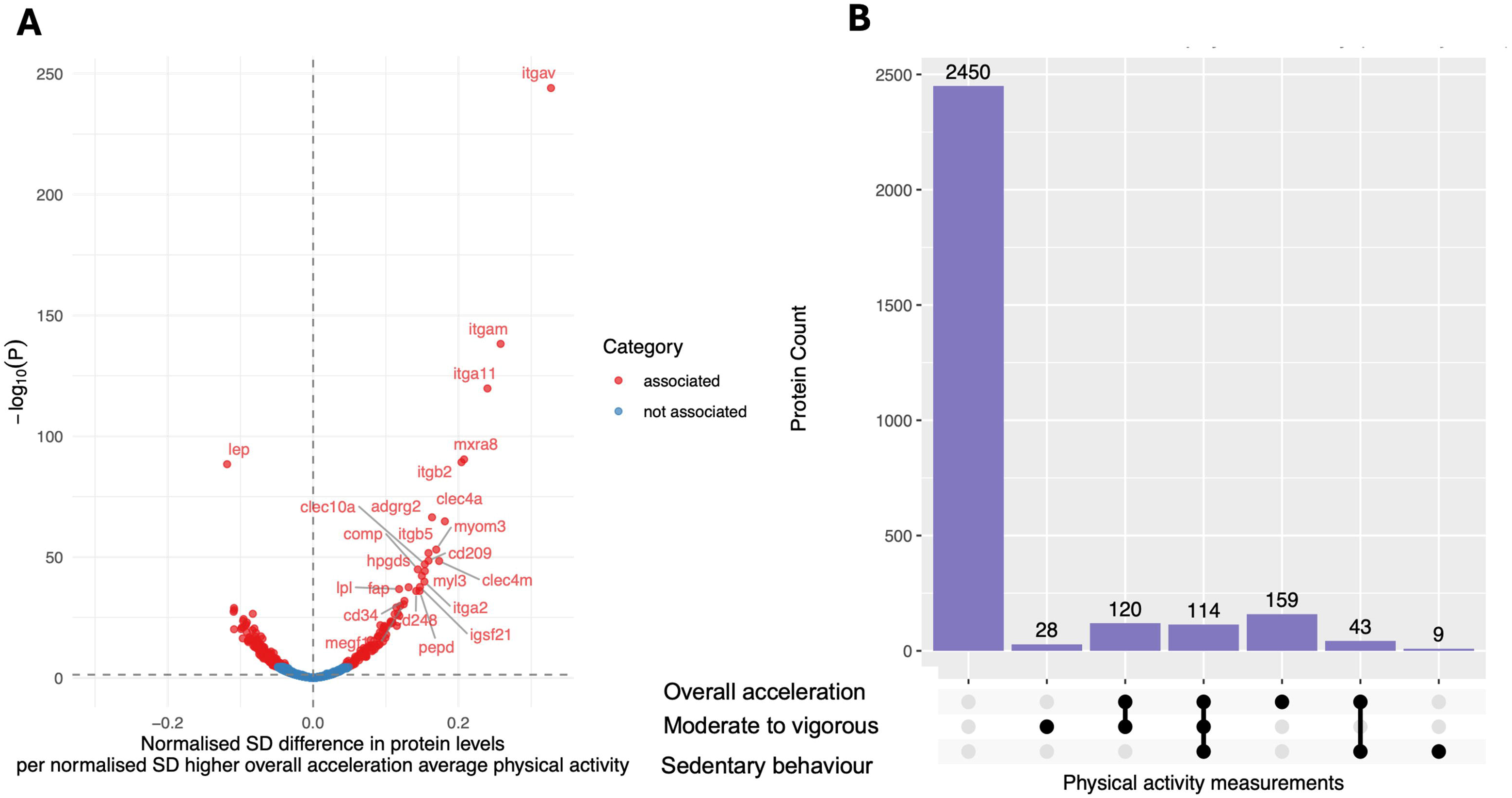

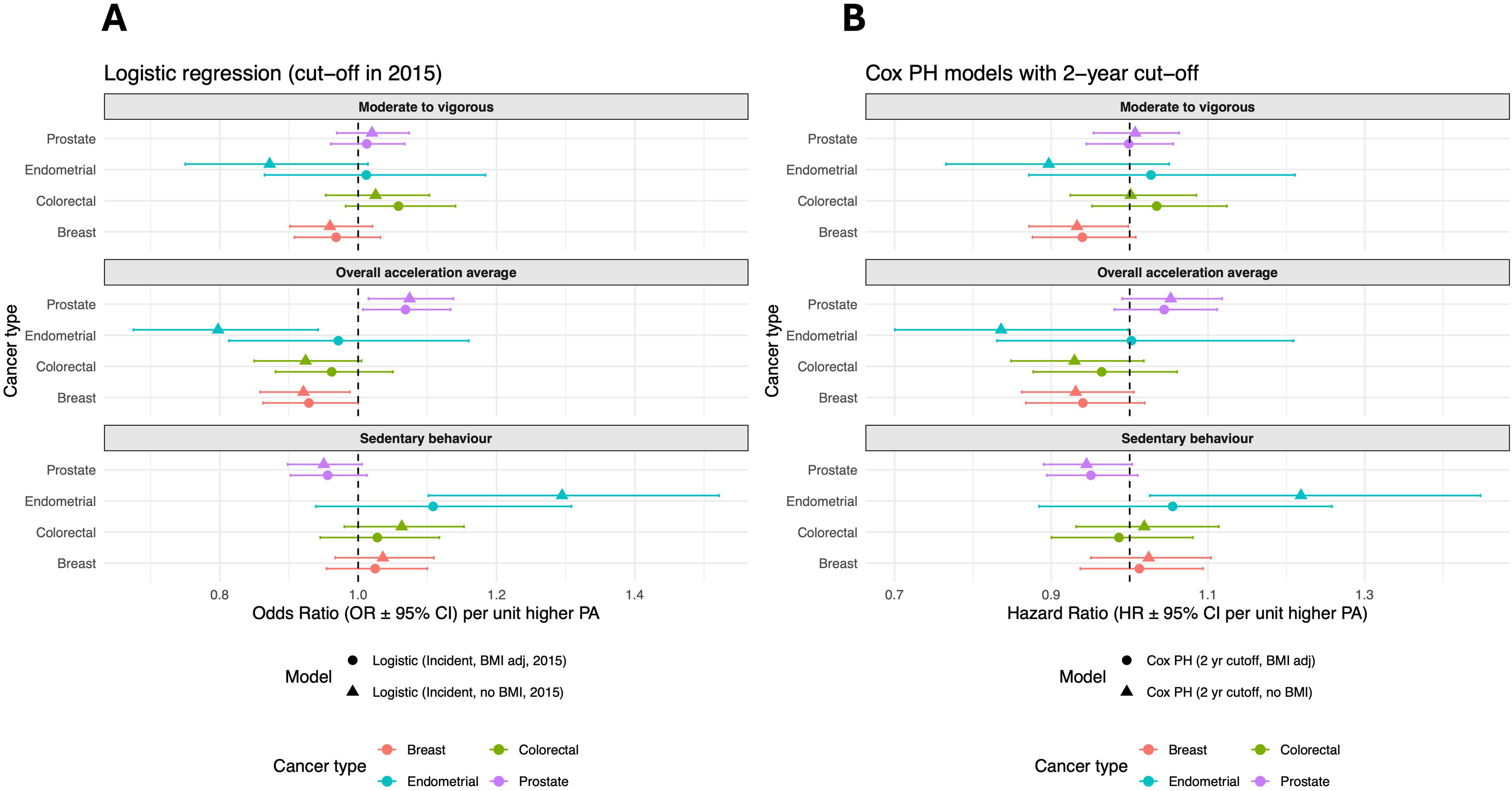

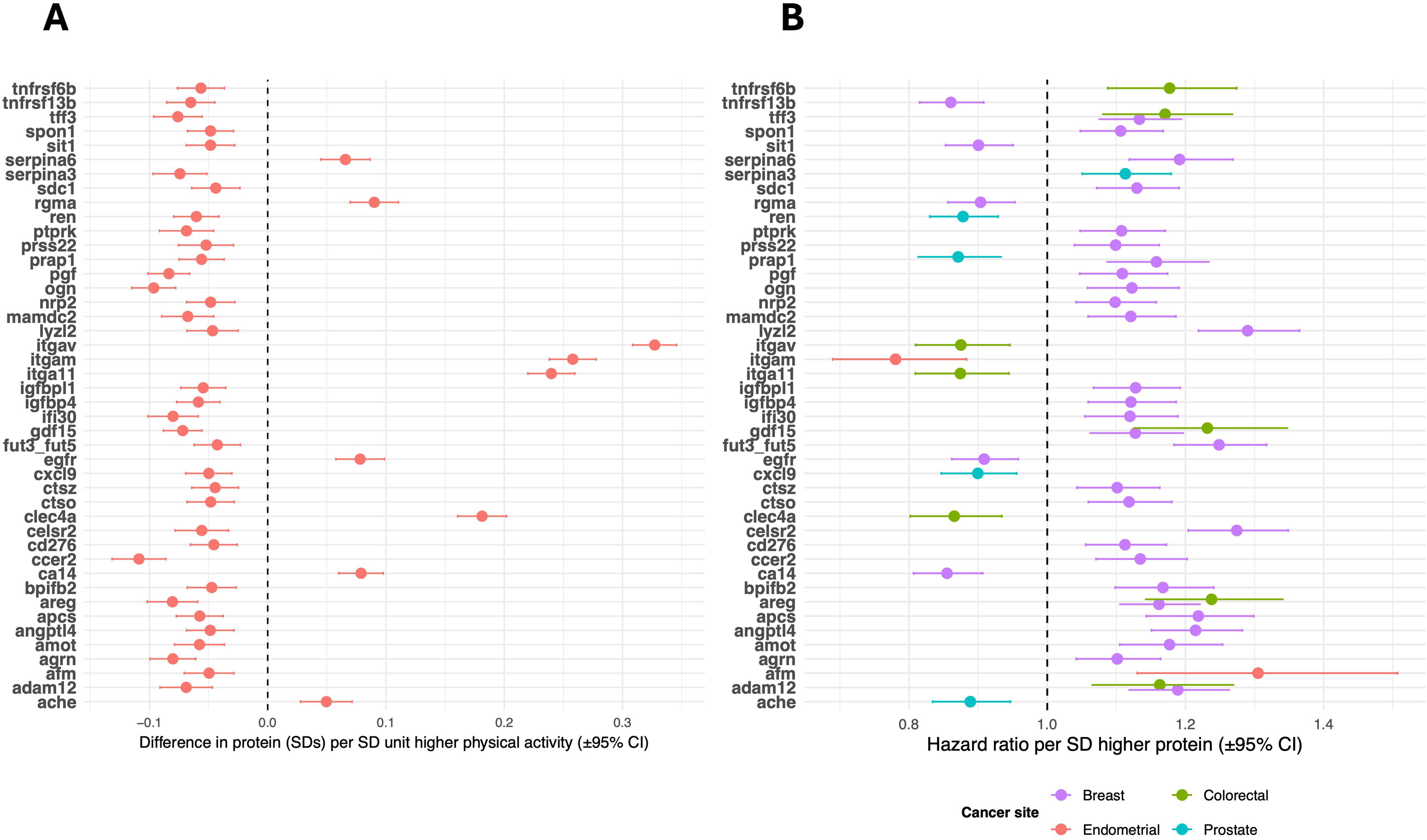

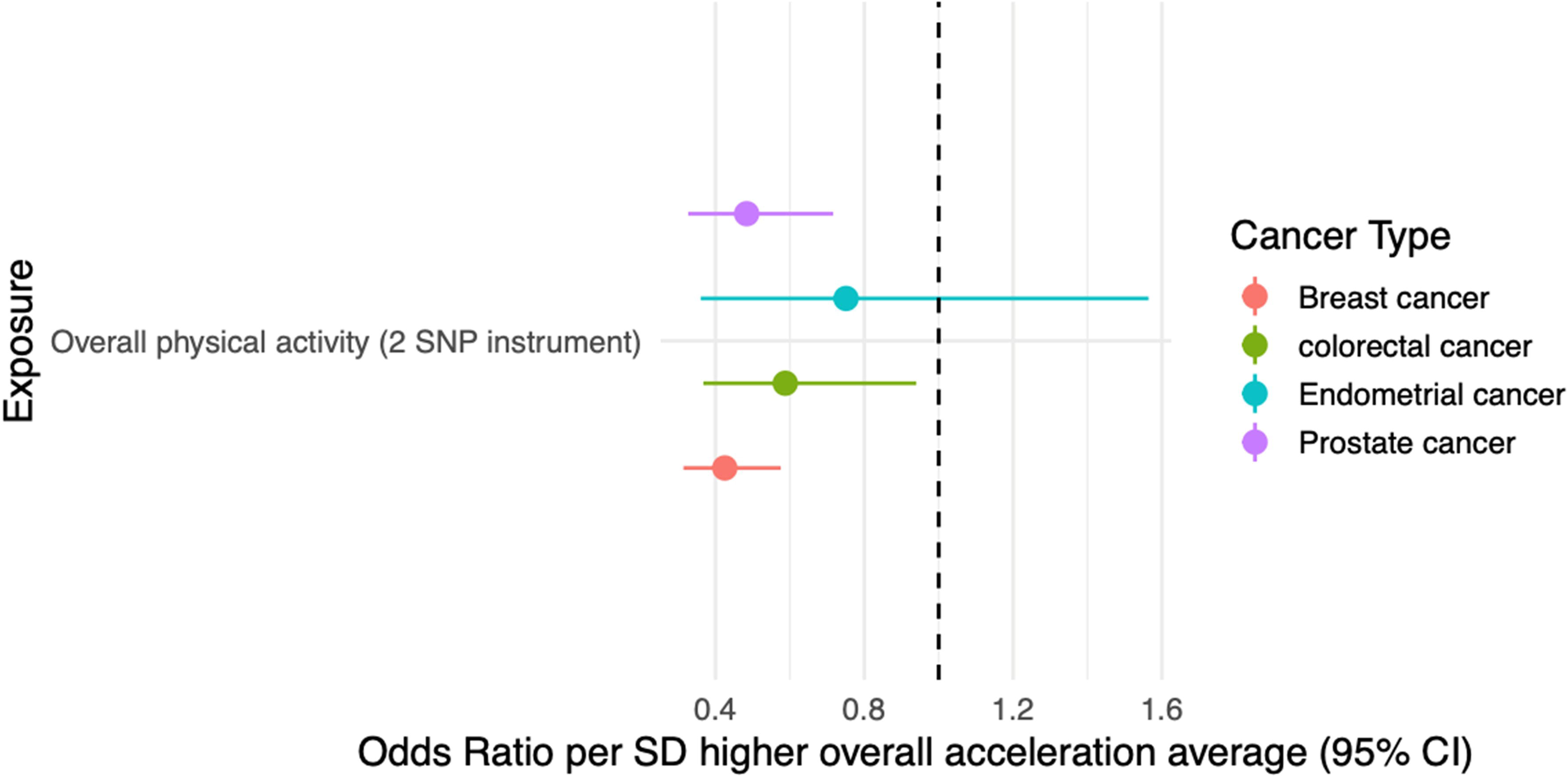

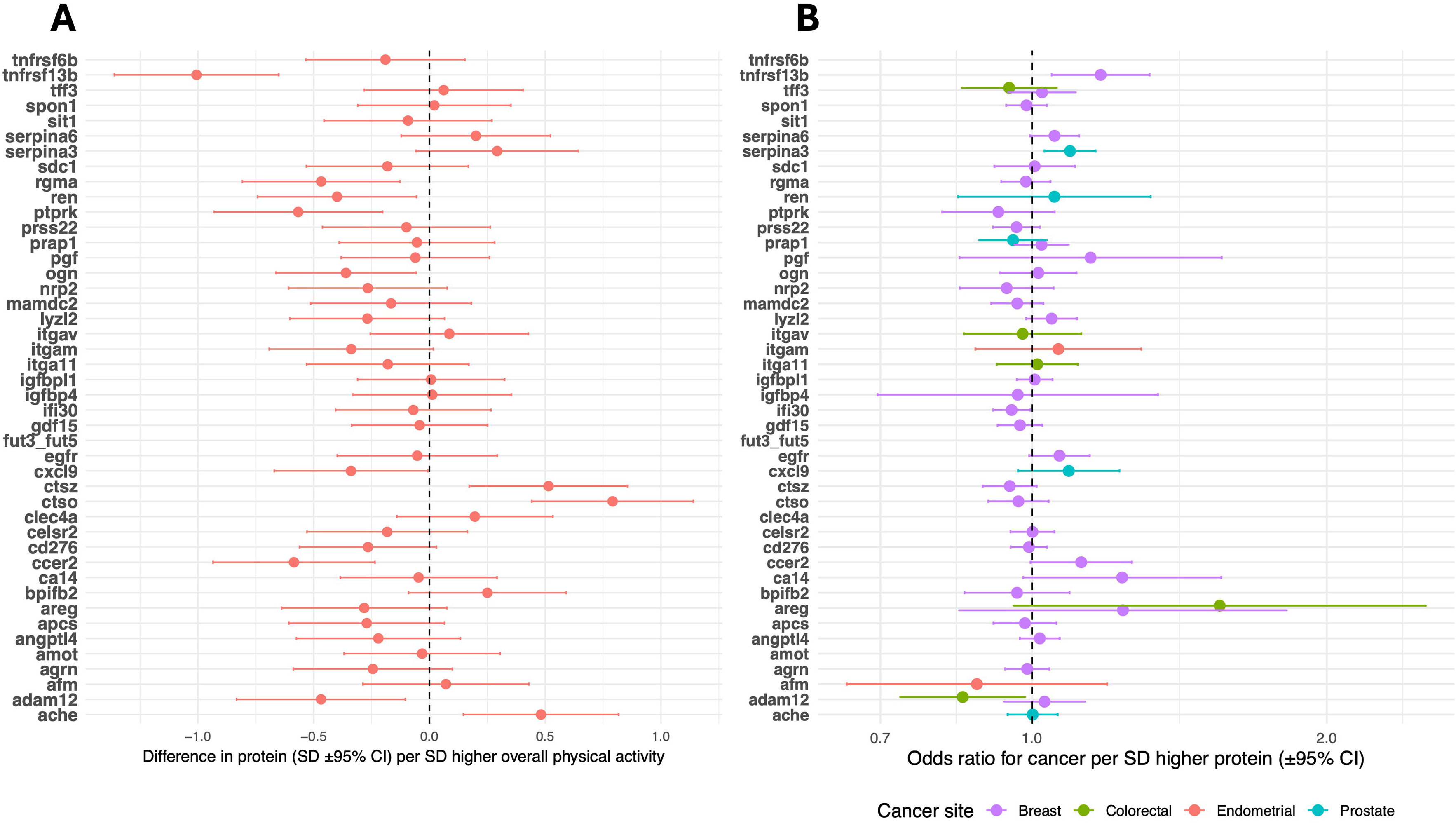

## Notes

### Competing Interest Statement

The authors have declared no competing interest.

