## Supplemental Methods for "Exploring proteins as intermediates between physical activity and cancer in UK biobank"

***Supplementary material***

*Cancer genome-wide association studies*

Genome-wide association study data were used for the MR analyses of colorectal, breast, prostate, endometrial cancers (1-4). Studies were restricted to those that included individuals of European ancestry. A brief summary of these GWAS is provided below, and full methodological details are available in the original publications.

For the prostate cancer GWAS, there were 79148 cases and 61106 controls (OpenGWAS ID: ieu-b-85) from the PRACTICAL consortium (1). Genotyping was performed using the Illumina OncoArray and iCOGS platforms. The OncoArray was developed by the NCI GAME-ON consortium and included around 79,000 prostate cancer-specific SNPs alongside a GWAs backbone (Illumina HumanCore) to enable high-resolution imputation. After QC, genotypes were imputed to the 1000 Genomes Project Phase 3 (October 2014, GRCh37) reference panel using IMPUTE2 for imputation, resulting in 20 million SNPs. Per allele ORs of prostate cancer were derived using a logistic regression, with adjustment for 7 principal components and covariates. OR estimates and SEs were combined by a fixed-effects inverse variance meta-analysis in METAL (5).

Breast cancer summary statistics were obtained from the Breast Cancer Association Consortium (BCAC) OncoArray meta-analysis (2) (Open GWAS ID: ieu-a-1126). This meta-analysis combined data from the OncoArray, iCOGS and 11 previous GWAS, totalling 122,977 cases and 105,974 controls. Cases were histologically confirmed as invasive breast cancer or ductal carcinoma in situ (DCIS) using cancer registries, medical records or pathology reports. Controls were women without a history of breast cancer. Genotyping was carried out using the Illumina OncoArray. Imputations to the 1000 Genomes Project Phase 3 reference panel was performed using SHAPEIT2 and IMPUTE2. Logistic regression was performed go generate each SNP OR, adjusting for country/study and the first 10 principal components. Fixed effects inverse-variance meta-analysis was implemented in METAL. In the publicly available summary statistics, there were ~10.7 million SNPs available.

GWAS summary statistics for endometrial cancer were generated with 12,906 cases and 108,979 controls 13 studies of endometrial cancer (4). Genotyping was conducted using the OncoArray, iCOGS and other GWAS platforms and imputations performed using SHAPEIT2 and IMPUTE2 with 1000 Genomes Project Phase 3 reference panel. There were approximately 11.7 million SNPs after QC and ~9.5 million in the publicly available dataset. Logistic regression was performed adjusted for genetic principal components, and fixed-effects inverse-variance (METAL) was use.

The colorectal cancer GWAS was performed using studies from the Genetics and Epidemiology of Colorectal Cancer Consortium (GECCO), the Colorectal Cancer Transdisciplinary Study (CORECT) and the Colon Cancer Family Registry (CCFR). The GWAS included 52,775 colorectal cancer cases and 45,940 controls and excluded individuals (3). CRC classification was determined using ICD-10 cases, with most cases being newly diagnosed. Genotyping was performed using multiple Illumina and Affymetrix arrays, and imputation was conducted using SHAPEIT2 for phasing and minimac3 with both a custom whole-genome sequence–based reference panel and the Haplotype Reference Consortium panel. Association analyses were performed using logistic regression adjusted for age, sex, study/genotyping project, and ancestry principal components, with results meta-analysed using fixed-effects inverse-variance weighting.
