## Supplementary material for "Exploring proteins as intermediates between physical activity and cancer in UK biobank": Checklist

**STROBE-MR checklist of recommended items to address in reports of Mendelian randomization studies**^1^ ^2^

| **Item No.** | **Section** | **Checklist item** | **Page No.** | **Relevant text from manuscript** |
| --- | --- | --- | --- | --- |
| 1 | **TITLE and ABSTRACT** | Indicate Mendelian randomization (MR) as the study’s design in the title and/or the abstract if that is a main purpose of the study | 1 | In abstract |
|  | **INTRODUCTION** |  |  |  |
| 2 | **Background** | Explain the scientific background and rationale for the reported study. What is the exposure? Is a potential causal relationship between exposure and outcome plausible? Justify why MR is a helpful method to address the study question | 2 | Higher levels of physical activity (PA) have consistently been shown to be associated with a reduced risk of several cancers…… the molecular intermediates which link PA to cancer risk are not fully understood. Characterising these biological pathways could inform strategies to optimise PA for cancer prevention and provide protein targets for potential modulation. Advances in high throughput proteomic profiling from small volumes of blood now offer opportunities to identify biological pathways influenced by PA that may underlie its protective effects on cancer. |
| 3 | **Objectives** | State specific objectives clearly, including pre-specified causal hypotheses (if any). State that MR is a method that, under specific assumptions, intends to estimate causal effects | 2-3 | This study used a combination of observational and MR approaches to further understand the associations between physical activity, circulating protein levels and cancer risk (**Figure 1**).  The main objectives were:   (1) A) Assess the associations between accelerometer-measured physical activity and circulating protein levels in the UK Biobank. B) Examine the relationships between physical activity and incident breast, prostate, endometrial, and colorectal cancers. C) Evaluate the associations between physical activity–related proteins and the risks of breast, prostate, endometrial, and colorectal cancer. (2) For proteins associated with overall acceleration average and cancer outcomes in a directionally concordant manner, apply two-step MR to investigate potential causal relationships. |
|  | **METHODS** |  |  |  |
| 4 | **Study design and data sources** | Present key elements of the study design early in the article. Consider including a table listing sources of data for all phases of the study. For each data source contributing to the analysis, describe the following: |  |  |
|  | a) | Setting: Describe the study design and the underlying population, if possible. Describe the setting, locations, and relevant dates, including periods of recruitment, exposure, follow-up, and data collection, when available. | 3, 6-7 | UK Biobank (UKB)  The UKB is a prospective study which recruited 500,000 participants across 22 assessment centres across the United Kingdom. At initial recruitment in 2006-2010, participants were aged 40-69 years [9]. This study has collected information about participants using a combination of questionnaires, physical and clinical measures [9]. UK Biobank also performs centralised linkage of participants to national hospital, cancer, and death registry records. |
|  | b) | Participants: Give the eligibility criteria, and the sources and methods of selection of participants. Report the sample size, and whether any power or sample size calculations were carried out prior to the main analysis | 6-7 | In sections describing PA, protein and cancer GWAS |
|  | c) | Describe measurement, quality control and selection of genetic variants | 6-7 | In methods section and supplementary materials |
|  | d) | For each exposure, outcome, and other relevant variables, describe methods of assessment and diagnostic criteria for diseases |  | In supplementary materials section |
|  | e) | Provide details of ethics committee approval and participant informed consent, if relevant | 12 | This study has received GECCO approval to use the colorectal cancer GWAS summary statistics file. |
| 5 | **Assumptions** | Explicitly state the three core IV assumptions for the main analysis (relevance, independence and exclusion restriction) as well assumptions for any additional or sensitivity analysis | 5 | This study employed a Mendelian randomization (MR) framework, which is based on three key instrumental variable assumptions. (i) Relevance: the selected genetic variants are strongly associated with the exposure. (ii) Independence: the genetic variants are not associated with confounding factors that influence both the exposure and the outcome. (iii) Exclusion restriction: the genetic variants affect the outcome exclusively through the exposure, with no alternative pathways (i.e., no horizontal pleiotropy). |
| 6 | **Statistical methods: main analysis** | Describe statistical methods and statistics used |  |  |
|  | a) | Describe how quantitative variables were handled in the analyses (i.e., scale, units, model) | 6-7 | The two overall physical activity GWAS were measured in different units: in the GWAS by Klimentidis et al. (main analysis), the GWAS betas were measured in milligravities, and for the GWAS by Doherty et al. (sensitivity analysis), the GWAS betas were converted to standard deviation (SD) units [18, 20]. For comparison with the observational analyses and with the analyses using the genetic instruments from the GWAS by Doherty et al., we converted the results from the MR analyses using the Klimentidis et al. GWAS to per SD units of physical activity, by multiplying the betas by the SD.  AND  Results are presented as an SD change (continuous) or odd ratio (dichotomous) per SD change in the exposure. |
|  | b) | Describe how genetic variants were handled in the analyses and, if applicable, how their weights were selected | 6-7 | In methods section |
|  | c) | Describe the MR estimator (e.g. two-stage least squares, Wald ratio) and related statistics. Detail the included covariates and, in case of two-sample MR, whether the same covariate set was used for adjustment in the two samples | 7 | Where ≥3 SNPs were available, an inverse variance weighted (IVW) method with random effects was used; where 2 or more SNPs were available, an IVW method with fixed effects was used; where 1 SNP was available, the Wald ratio was used [29-31]. |
|  | d) | Explain how missing data were addressed |  | NA |
|  | e) | If applicable, indicate how multiple testing was addressed | 5 | Proteins included in these analyses were brought forward from observational analyses, such that proteins which had evidence for an association with overall physical activity and with any of the cancer outcomes in fully adjusted models were included, therefore for these analyses we were looking for consistency with the observational analyses rather than applying strict Bonferroni corrections. |
| 7 | **Assessment of assumptions** | Describe any methods or prior knowledge used to assess the assumptions or justify their validity | 7 | Sensitivity analyses (where ≥3 SNPs were available) included weighted median, weighted mode and MR Egger, which were applied to assess the robustness of the causal estimates and to detect potential violations of MR exclusion assumption, which can be caused by horizontal pleiotropy [32-34]. Heterogeneity was explored through use of Cochran’s Q statistic [35]. To assess the relevance assumption we calculated instrument F-statistics, to assess independence we reported other phenotypes associated with our instruments based on data from GWAS catalog <https://www.ebi.ac.uk/gwas/>. |
| 8 | **Sensitivity analyses and additional analyses** | Describe any sensitivity analyses or additional analyses performed (e.g. comparison of effect estimates from different approaches, independent replication, bias analytic techniques, validation of instruments, simulations) | 7 | See above |
| 9 | **Software and pre-registration** |  |  |  |
|  | a) | Name statistical software and package(s), including version and settings used | 7 | Two-sample Mendelian randomization was performed using the “TwoSampleMR” R package. |
|  | b) | State whether the study protocol and details were pre-registered (as well as when and where) |  | NA |
|  | **RESULTS** |  |  |  |
| 10 | **Descriptive data** |  |  |  |
|  | a) | Report the numbers of individuals at each stage of included studies and reasons for exclusion. Consider use of a flow diagram |  | Reported in supplementary tables |
|  | b) | Report summary statistics for phenotypic exposure(s), outcome(s), and other relevant variables (e.g. means, SDs, proportions) | 17 | Table 1 |
|  | c) | If the data sources include meta-analyses of previous studies, provide the assessments of heterogeneity across these studies |  | NA |
|  | d) | For two-sample MR:  i.  Provide justification of the similarity of the genetic variant-exposure associations between the exposure and outcome samples  ii.  Provide information on the number of individuals who overlap between the exposure and outcome studies | 7 | Some overlap existed between the GWAS used for the exposures and the outcome, for physical activity and circulating proteins, both of which included UK Biobank participants, the degree of sample overlap was limited; For example, based on exploring overlap of the individual level data in UKB, approximately 20% of participants in the protein GWAS were also included in the physical activity GWAS, and about 12% of the physical activity GWAS sample overlapped with the protein GWAS sample [36]. |
| 11 | **Main results** |  |  |  |
|  | a) | Report the associations between genetic variant and exposure, and between genetic variant and outcome, preferably on an interpretable scale |  | Supplementary Table 13 |
|  | b) | Report MR estimates of the relationship between exposure and outcome, and the measures of uncertainty from the MR analysis, on an interpretable scale, such as odds ratio or relative risk per SD difference |  | Supplementary Tables 14-19 |
|  | c) | If relevant, consider translating estimates of relative risk into absolute risk for a meaningful time period |  | NA |
|  | d) | Consider plots to visualize results (e.g. forest plot, scatterplot of associations between genetic variants and outcome versus between genetic variants and exposure) |  | Figs 5 and 6 |
| 12 | **Assessment of assumptions** |  |  |  |
|  | a) | Report the assessment of the validity of the assumptions |  |  |
|  | b) | Report any additional statistics (e.g., assessments of heterogeneity across genetic variants, such as *I^2^*, Q statistic or E-value) |  | Supplementary Table 20 |
| 13 | **Sensitivity analyses and additional analyses** |  |  |  |
|  | a) | Report any sensitivity analyses to assess the robustness of the main results to violations of the assumptions |  | Supplementary Table 14, 15 and 19 |
|  | b) | Report results from other sensitivity analyses or additional analyses |  | NA |
|  | c) | Report any assessment of direction of causal relationship (e.g., bidirectional MR) |  | Supplementary Table 15 and 19 |
|  | d) | When relevant, report and compare with estimates from non-MR analyses | 8-9 | Observational analyses are the primary analyses in this study |
|  | e) | Consider additional plots to visualize results (e.g., leave-one-out analyses) |  |  |
|  | **DISCUSSION** |  |  |  |
| 14 | **Key results** | Summarize key results with reference to study objectives | 10-12 | In discussion |
| 15 | **Limitations** | Discuss limitations of the study, taking into account the validity of the IV assumptions, other sources of potential bias, and imprecision. Discuss both direction and magnitude of any potential bias and any efforts to address them | 12-13 | In discussion |
| 16 | **Interpretation** |  |  |  |
|  | a) | Meaning: Give a cautious overall interpretation of results in the context of their limitations and in comparison with other studies | 11-13 | In discussion |
|  | b) | Mechanism: Discuss underlying biological mechanisms that could drive a potential causal relationship between the investigated exposure and the outcome, and whether the gene-environment equivalence assumption is reasonable. Use causal language carefully, clarifying that IV estimates may provide causal effects only under certain assumptions | 11-13 | In discussion |
|  | c) | Clinical relevance: Discuss whether the results have clinical or public policy relevance, and to what extent they inform effect sizes of possible interventions | 13 | Our study shows robust evidence for a protective effect of physical activity on cancer risk and adds to the body of evidence on this. |
| 17 | **Generalizability** | Discuss the generalizability of the study results (a) to other populations, (b) across other exposure periods/timings, and (c) across other levels of exposure |  | This will also affect the generalisability of our findings to other populations which are not primarily white European. Similarly, we assumed linear models and were looking at the effects of exposures within the normal range we were unable to examine the effects of exposures at the extreme end of the spectrum, nor where we able to assess the importance of timing of exposure in this study. |
|  | **OTHER INFORMATION** |  |  |  |
| 18 | **Funding** | Describe sources of funding and the role of funders in the present study and, if applicable, sources of funding for the databases and original study or studies on which the present study is based | 13 | Included |
| 19 | **Data and data sharing** | Provide the data used to perform all analyses or report where and how the data can be accessed, and reference these sources in the article. Provide the statistical code needed to reproduce the results in the article, or report whether the code is publicly accessible and if so, where | 13 | Scripts used for analyses in this manuscript are located here: <https://github.com/lucygoudswaard/PA_protein_cancer_paper>. ChatGPT was used to support with analytical code writing. |
| 20 | **Conflicts of Interest** | All authors should declare all potential conflicts of interest | 13 | All authors declare no conflicts of interest. |

This checklist is copyrighted by the Equator Network under the Creative Commons Attribution 3.0 Unported (CC BY 3.0) license.

1.	Skrivankova VW, Richmond RC, Woolf BAR, Yarmolinsky J, Davies NM, Swanson SA, et al. Strengthening the Reporting of Observational Studies in Epidemiology using Mendelian Randomization (STROBE-MR) Statement. JAMA. 2021;under review.

2.	Skrivankova VW, Richmond RC, Woolf BAR, Davies NM, Swanson SA, VanderWeele TJ, et al. Strengthening the Reporting of Observational Studies in Epidemiology using Mendelian Randomisation (STROBE-MR): Explanation and Elaboration. BMJ. 2021;375:n2233.
